## Supplementary Material for "Development and preliminary testing of Health Equity Across the AI Lifecycle (HEAAL): A framework for healthcare delivery organizations to mitigate the risk of AI solutions worsening health inequities"

### Supporting Information

**S1 Appendix. Overview of Health Equity Across the AI Lifecycle (HEAAL).** For evaluating an existing AI solution, follow procedures written in red and black text. For evaluating a new AI solution, follow procedures written in blue and black text.

| Adoption stage | Decision point | Assessment domains | Procedures |
|---|---|---|---|
| Problem identification and procurement | 1. Identify and prioritize a problem | Fitness for purpose | a. Ensure that problems are prioritized and funded equally across all patient subgroups.<br>b. Determine whether there are patient populations for whom a solution to the prioritized problem should not be used, should be used differently, or whose experience with the system should be closely monitored. |
| | 2. Define AI product scope and intended use | Reliability and validity, Fairness | a. List alternative solutions for the problem, including non-technical interventions and other non-AI technical interventions.<br>b. Define an ideal label for model development.<br>c. Seek an approval from an institutional review board, ethical review board, or research ethics board to access and use local healthcare retrospective data.<br>d. Assess health inequities present in the local healthcare retrospective data and identify disadvantaged patient subgroups within the context of the prioritized problem.<br>e. Examine whether a local healthcare retrospective data set is representative of demographic representation of local non-healthcare data.<br>f. Assess health inequities present in the model training data and identify disadvantaged patient subgroups within the context of the prioritized problem.<br>g. Examine whether the model training data is representative of the demographics present within the local healthcare retrospective data.<br>h. Analyze label choice bias across disadvantaged and advantaged patient subgroups.<br>i. Ensure that the model features are relevant to its actual label and capture the same meanings across disadvantaged and advantaged patient subgroups.<br>j. Identify potential hidden stratification that masks unequal model performance between disadvantaged and advantaged patient subgroups.<br>k. Gather model performance data and compare it between disadvantaged and advantaged patient subgroups.<br>l. Determine which SDOH and demographic data are appropriate to be included in the model to minimize potential risk of worsening health inequities.<br>m. Determine which potential solution best solves the problem for disadvantaged patient subgroups. |

| | | | |
|---|---|---|---|
| Development and adaptation | 3. Develop success measures | Fairness | a. Establish equity objectives for implementation of the selected AI solution.<br>b. Identify the most appropriate fairness metrics to use for the selected AI product and its design. |
| | 4. Design AI solution workflow | Fairness, Fitness for purpose, Transparency | a. Ensure that the solution design is informed by meaningful and pragmatic recommendations from members of disadvantaged patient subgroups.<br>b. Ensure that the solution design promotes inclusivity of clinical end-users and usability of the solution.<br>c. Design complementary non-technical solution components required to achieve equity objectives for implementation of the solution.<br>d. Align clinical end-users and organizational leaders to achieve equity objectives. |
| | 5. Generate evidence of safety, efficacy and equity | Accountability, Fairness, Reliability and validity | a. Assess completeness and quality of local data required to construct model features across disadvantaged patient subgroups.<br>b. Seek an approval from an institutional review board, ethical review board, or research ethics board to access and use local healthcare prospective data.<br>c. Analyze label choice bias across disadvantaged and advantaged patient subgroups.<br>d. Ensure that the model features are relevant to its actual label and capture the same meanings across disadvantaged and advantaged patient subgroups.<br>e. Identify potential hidden stratification that masks unequal model performance between disadvantaged and advantaged patient subgroups.<br>f. Assess model performance and compare it between disadvantaged and advantaged patient subgroups.<br>g. Determine which SDOH and demographic data are appropriate to be included in the model to minimize potential risk of worsening health inequities.<br>h. Assess prospective model performance and compare it to retrospective model performance across disadvantaged and advantaged patient subgroups.<br>i. Perform comprehensive assessment of model performance across disadvantaged and advantaged patient subgroups.<br>j. If the model performs worse for a certain patient subgroup on local prospective healthcare data, consider adapting the model or its use to help minimize negative impacts on inequities.<br>k. Ensure that model performance aligns with the equity objectives.<br>l. Conduct a prospective pilot study to validate whether the AI solution achieves equity objectives. |

| | | | |
|---|---|---|---|
| Clinical integration | 6. Execute AI solution rollout | Accountability, Fairness, Transparency | a. Document information about the model development and implementation and share it with clinical end-users, members of disadvantaged patient subgroups, and others who may be affected by use of the model.<br>b. Educate clinical end-users about potential bias in using the solution.<br>c. Where applicable, seek an approval from an institutional review board, ethical review board, or research ethics board to implement and use the AI solution in clinical practice.<br>d. After rollout, continue to seek feedback from clinical end-users and members of disadvantaged and advantaged patient subgroups to achieve equity objectives. |
| Lifecycle management | 7. Monitor the AI solution | Fairness, Reliability and validity, Transparency | a. Regularly monitor the model performance across disadvantaged and advantaged patient subgroups.<br>b. Regularly monitor the work environment of the solution across disadvantaged and advantaged patient subgroups.<br>c. Regularly monitor health inequities across disadvantaged and advantaged patient subgroups. |
| | 8. Update or decommission the AI solution | Accountability, Fairness, Reliability and validity, Transparency | a. Determine updates to the model or its work environment.<br>b. If updating the model or its work environment does not improve model performance or fails to improve progress towards equity objectives, consider decommissioning the model.<br>c. If the model successfully achieved equity objectives and there is interest to expand model use, evaluate appropriate technical and non-technical resources to expand the AI solution to new settings. |

**S2 Appendix. Health Equity Across the AI Lifecycle (HEAAL).**

### 1. Identify and prioritize a problem

[Assessment domain: Fitness for purpose]

Decision Point 1 focuses on identifying and prioritizing problems within health care delivery organizations and surfacing stakeholders affected by problems. It also describes how to determine different dimensions of problems and assess the suitability of AI as a technical approach to address problems. Decision Point 1 starts with surfacing problems across the organization and ends with a prioritized problem to solve.

a. Ensure that problems are prioritized and funded equally across all patient subgroups. [Active stakeholders: S]
    i. Prioritize problems that demonstrate potential to positively impact all patient subgroups.
    ii. Ensure to account for the number and types of affected patients in making funding and resource allocation decisions.

b. Determine whether there are patient populations for whom a solution to the prioritized problem should not be used, should be used differently, or whose experience with the system should be closely monitored.
[Active stakeholders: C, O, P] [Data source: Literature review, Qualitative data]
    i. Review literature on epidemiology and health disparities to understand the current state of health inequities within the context of the prioritized problem.[1]
    ii. Identify social determinants of health (SDOH) and demographic subgroups who may experience health inequities (i.e., disadvantaged patient subgroups) within the context of the prioritized problem.[2]
    iii. Continue identifying health inequities and disadvantaged patient subgroups by interviewing or surveying personnel and patient community members best positioned to understand the experiences of disadvantaged patient subgroups.[3]
    iv. Drawing from information collected from the previous procedures, establish a list of health inequities and disadvantaged patient subgroups. Make sure to list

[1] Understanding the current state is important for ensuring that the proposed solution accounts for contextual characteristics of patients that may shape the inequities so that all patients can receive equitable care. Review Table 2 for examples of different types of health inequities.

[2] It is important to note that the absence of documented inequalities does not mean the absence of inequalities. For example, some demographic subgroups, such as transgender and gender-nonconforming individuals and undocumented immigrants, have poor data collection to even understand inequities. (Chen IY, Pierson E, Rose S, Joshi S, Ferryman K, Ghassemi M. Ethical Machine Learning in Healthcare. Annu Rev Biomed Data Sci. 2021;4(1):1-22. doi:10.1146/annurev-biodatasci-092820-114757) While information about language that patients speak may not have been captured, language serves as an important social identity of patients.

[3] Such personnel may include healthcare providers, social workers, patient navigators, and patient community members identified as negatively impacted by health inequities. Sample questions to ask the personnel are described in the worksheet.

intersecting identities that may be associated with worse inequities and include references.

#### 2. Define AI product scope and intended use

[Assessment domain: Fairness, Reliability and validity]

Decision Point 2 describes how organizations can assess the feasibility and viability of adopting AI to solve problems. It also explains how to conduct preliminary assessments of AI products, assess legal risks of AI product adoption, and audit the process by which AI product investments are made. Decision Point 2 starts with the assessment of internal capability and ends with a decision to buy or build an AI product.

a. List alternative solutions for the problem, including non-technical interventions and other non-AI technical interventions.
   [Active stakeholders: C, O, T]

b. Define an ideal label for model development.
   [Active stakeholders: C, O, P, T] [Data source: Local healthcare retrospective data, Qualitative data]
   i. Conduct interviews, focus groups, or surveys with a cross-functional team to examine how patients should be identified by an algorithm to optimize decision making within the context of the prioritized problem. Allow the cross-functional team to assume that the algorithm had access to pristine and complete information about all patients.[4]
   ii. Assess responses from the cross-functional team and identify the most promising clinical outcome used for model development which will be defined as an ideal label. Ensure to document the rationale for selecting the label.

c. Seek an approval from an institutional review board, ethical review board, or research ethics board to access and use local healthcare retrospective data.
   [Active stakeholders: R]

d. Assess health inequities present in the local healthcare retrospective data and identify disadvantaged patient subgroups within the context of the prioritized problem.
   [Active stakeholders: T] [Data source: Local healthcare retrospective data]
   i. Specify which data elements can be used to measure health inequities identified in 1(b)(iv).
   ii. Using the identified data elements, examine whether health inequities identified in 1(b)(iv) are present within local healthcare retrospective data.

---

[4] Individuals and groups who understand the experiences of disadvantaged patient subgroups can provide important context around the labels and suggest a desirable state that could be pursued using the algorithm. (Mccradden M, Odusi O, Joshi S, et al. What’s fair is… fair? Presenting JustEFAB, an ethical framework for operationalizing medical ethics and social justice in the integration of clinical machine learning. 2023 ACM Conf Fairness, Account, Transpar. Published online 2023:1505-1519. doi:10.1145/3593013.3594096)

iii. Combine information collected from the previous procedure 2(d)(ii) with information collected from 1(b)(iv) to establish a list of health inequities and disadvantaged patient subgroups within the local healthcare delivery setting.[5] The list should include references and supporting data. All other patient subgroups, not including the disadvantaged patient subgroups, are defined as advantaged patient subgroups.[6]

e. Examine whether a local healthcare retrospective data set is representative of demographic representation of local non-healthcare data.
[Active stakeholders: T] [Data source: Local healthcare retrospective data, Local non-healthcare data]
   i. Conduct a demographic analysis of the patient cohort within the local healthcare retrospective data set.
   ii. Examine whether the demographics of the local healthcare retrospective data reflects demographic representation of local non-healthcare data derived from external data sources.[7]
   iii. If demographics of the local healthcare retrospective data does not significantly align with demographics of the local non-healthcare data, flag for potential *representation bias*.

**If considering an existing AI solution (regardless of whether it was developed internally or externally), proceed to the next set of procedures written in red to assess AI solution options and select the most optimal one. If an AI solution does not exist and is being considered for internal development, skip to Decision Point 3.**

f. Assess health inequities present in the model training data and identify disadvantaged patient subgroups within the context of the prioritized problem.
[Active stakeholders: T] [Data source: Training data]
   i. Using data elements that are similar to the ones identified in 2(d)(i), examine whether health inequities identified in 1(b)(iv) are present within the training data.
   ii. Confirm the presence or absence of health inequities among disadvantaged patient subgroups previously identified in 2(d)(iii).
   iii. The implication of the presence or absence of health inequities within the training data depends on whether health inequities were identified within local healthcare retrospective data in 2(d)(ii):
      - If health inequities were present within local healthcare retrospective data, the presence or absence of health inequities within the training data conveys little to no concern for worsening existing health inequities in the local setting.
      - If health inequities were not present within local healthcare retrospective data, the absence of health inequities within the training data conveys

[5] For example, if a patient subgroup A was identified as a disadvantaged patient subgroup in 1(b)(iv) and a patient subgroup B was identified as a disadvantaged patient subgroup in 2(d)(ii), both patient subgroups A and B should be considered as disadvantaged patient subgroups.
[6] Keep in mind that the list of disadvantaged patient subgroups and advantaged patient subgroups may be updated as the analysis progresses.
[7] For example, US Census demographic information can be used to curate demographic information about the population within a catchment area of a healthcare delivery organization.

little concern for creating future health inequities. However, the presence of health inequities within the training data conveys potential concern for creating future health inequities.

g. Examine whether the model training data is representative of the demographics present within the local healthcare retrospective data.
[Active stakeholders: T] [Data source: Training data]
   i. For each AI solution, conduct a demographic analysis of the patient cohort within the training data.
   ii. Examine whether the training data reflects a demographic representation of the local healthcare retrospective data analyzed in 2(e)(i).[8]
   iii. If demographics of the training data does not significantly align with demographics of the local healthcare retrospective data, flag for potential *representation bias*. Prioritize AI solutions that are trained on data sets that are representative of the patient subgroups within the local healthcare context.

h. Analyze label choice bias across disadvantaged and advantaged patient subgroups.
[Active stakeholders: C, O, P, T] [Data source: Training data]
   i. For each AI solution, gather a list of the actual labels used for model training.
   ii. From the list, select a closer-to-ideal label, even if it is available only for a small subset of patients. Use this label for analysis in the next procedures.
   iii. Consult with a cross-functional team and assess how well the actual label aligns with the ideal label identified in 2(b)(ii) across disadvantaged and advantaged patient subgroups identified in 2(d)(iii).
   iv. If the actual label does not accurately capture the ideal label, flag for *label choice bias*. Prioritize AI solutions with actual labels that accurately measure the ideal label.

i. Ensure that the model features are relevant to its actual label and capture the same meanings across disadvantaged and advantaged patient subgroups.
[Active stakeholders: C, T] [Data source: Training data]
   i. For each AI solution, gather the actual labels and a comprehensive list of model features used for model training.
   ii. Ensure that all model features accurately represent concepts relevant to the actual label.
   iii. Consult with clinicians and understand if there are known differences in how model features are gathered and represented across disadvantaged and advantaged patient subgroups identified in 2(d)(iii).[9]
   iv. For each feature, examine how it is correlated with actual labels across all patient subgroups and identify whether this association manifests inconsistently in disadvantaged patient subgroups identified in 2(d)(iii).

---

[8] For example, if there is a minimal representation of patient subgroups identified in section 1(b)(iv) in the training data, this representation should be a big red flag.
[9] For example, examine whether there are any known symptoms that manifest differently for different patient demographics.

v. If the association between model features and the actual label is inconsistent across different patient subgroups, flag for *measurement bias*. Prioritize AI solutions that use features that have consistent meanings and are captured robustly across all patient subgroups.

j. Identify potential hidden stratification that masks unequal model performance between disadvantaged and advantaged patient subgroups.
[Active stakeholders: T] [Data source: Training data]
    i. Identify diagnostic and treatment subgroups within the actual label.[10]
    ii. Compare model performance between the identified subgroups.
    iii. If the model performance of a subgroup with diagnosis and treatment is better than the model performance of a subgroup without diagnosis and treatment, flag for *hidden stratification*.[11] Consider alternative solutions (return to 2(a)) or retrain the model after excluding a cohort of patients from the subgroup with diagnosis and treatment.
    iv. If the model performance of a subgroup without diagnosis and treatment is better than the model performance of a subgroup with diagnosis and treatment, or if model performance between the subgroups is similar, then compare model performance between disadvantaged and advantaged patient subgroups defined in 2(d)(iii) within each diagnostic and treatment subgroup.
    v. Prioritize AI solutions that perform equally well on disadvantaged and advantaged patient subgroups within each diagnostic and treatment subgroup.

k. Gather model performance data and compare it between disadvantaged and advantaged patient subgroups.
[Active stakeholders: T] [Data source: Training data]
    i. For each AI solution, gather model performance data for disadvantaged patient subgroups and advantaged patient subgroups identified in 2(d)(iii).
    ii. Compare the model performance of disadvantaged patient subgroups to the model performance of advantaged patient subgroups.
    iii. If model performance is worse for disadvantaged patient subgroups than model performance for advantaged patient subgroups, the implication depends on whether health inequities were identified in 2(d)(ii):
        ▪ If health inequities were confirmed, the difference in model performance could be due to existing health inequities, not due to modeling, and thus, conveys minimal concern with the model.

---

[10] For example, diagnostic and treatment subgroups may include (1) a subgroup that has completed all relevant prior workup versus a subgroup who has not (workup inequity), (2) a subgroup that is diagnosed at an earlier disease state versus a subgroup that is diagnosed at a later disease state (diagnosis inequity), (3) a subgroup that receives important interventions prior to or immediately after diagnosis versus a subgroup that does not receive important interventions (treatment inequity), and (4) a subgroup with poor outcomes versus a subgroup with good outcomes (outcome inequity).
[11] Oakden-Rayner L, Dunnmon J, Carneiro G, Ré C. Hidden stratification causes clinically meaningful failures in machine learning for medical imaging. In: Vol 52. ACM CHIL '20: ACM Conference on Health, Inference, and Learning. ; 2020:151-159. doi:10.1145/3368555.3384468

- If health inequities were not confirmed, the difference in model performance could be due to modeling and create future health inequities and thus, conveys potential concern with the model.

iv. If model performance for disadvantaged patient subgroups is similar to model performance for advantaged subgroups, the implication depends on whether health inequities were identified in 2(d)(ii):
   - If health inequities were confirmed, the similarity in model performance could be due to modeling and further reinforce existing health inequities and thus, conveys potential concern with the model.
   - If health inequities were not confirmed, the similarity in model performance could be due to the absence of health inequities and thus, conveys minimal concern with the model.

v. Prioritize AI solutions with minimal concern.[12]

l. Determine which SDOH and demographic data are appropriate to be included in the model to minimize potential risk of worsening health inequities.
[Active stakeholders: T] [Data source: Training data]
   i. For each AI solution, review the comprehensive list of model features gathered in 2(h)(i).
   ii. Identify solutions that use patient SDOH and demographic data as model features.
   iii. For solutions that use patient SDOH and demographic data as model features, gather a rationale for inclusion of each SDOH and demographic feature.
   iv. Gather model performance data without using SDOH and demographic data as model features.
   v. Compare model performance data gathered in the previous procedure 2(m)(iv) with model performance data with SDOH and demographic features gathered in 2(k)(i) across disadvantaged and advantaged patient subgroups identified in 2(d)(iii).[13]
   vi. Prioritize AI solutions that use SDOH and demographic data to improve performance for disadvantaged patient subgroups identified in 2(d)(iii). However, if model performance remains robust across disadvantaged patient subgroups without using SDOH and demographic data, minimize use of this data.

m. Determine which potential solution best solves the problem for disadvantaged patient subgroups.
[Active stakeholders: C, O, T, S]

---

[12] For patient subgroups identified in section 2(d)(iii) who are poorly represented in the local context (e.g., Native American women in Boston), AI solution performance may have wide confidence bounds. Ensure that performance measures account for uncertainty.

[13] Sometimes, removing SDOH and demographic data as model features may decrease model performance. For example, removing race worsened model performance (Khor S, Haupt EC, Hahn EE, Lyons LJL, Shankaran V, Bansal A. Racial and Ethnic Bias in Risk Prediction Models for Colorectal Cancer Recurrence When Race and Ethnicity Are Omitted as Predictors. JAMA Netw Open. 2023;6(6):e2318495. doi:10.1001/jamanetworkopen.2023.18495).

i. Select a solution, which may or may not use AI, that best addresses the prioritized problem for disadvantaged patient subgroups identified in 2(d)(iii).
ii. If an AI solution is selected, proceed to Decision Point 3 and onward.

### 3. Develop success measures

[Assessment domain: Fairness]

Decision Point 3 focuses on defining the scope of use, constraints, and dependencies of AI solutions. It also describes how to define technical performance targets for AI products as well as measures of success for AI products used in practice. Decision Point 3 starts with development or local adaptation of the selected AI solution and ends with definition of success measures for its use.

a. Establish equity objectives for implementation of the selected AI solution.
   [Active stakeholders: C] [Data source: Organizational data]
   i. Review baseline inequities calculated in 2(d)(ii).
   ii. For each disadvantaged and advantaged patient subgroup identified in 2(d)(iii), set objectives for AI solution implementation to address health inequities identified in 2(d)(ii) in terms of health and economic outcomes.[14] Make sure to establish a cut-off or threshold for considering when to decommission the solution.

b. Identify the most appropriate fairness metrics to use for the selected AI product and its design.
   [Active stakeholders: C, O, T] [Data source: Literature review]
   i. Review the context of the use case and disadvantaged patient subgroups identified in 2(d)(iii).
   ii. Review literature and identify fairness metrics relevant to the solution.
   iii. Discuss which fairness metrics can be used to achieve equity objectives defined in 3(a)(ii).
   iv. Discuss which fairness metrics are pragmatic and align with health system priorities.
   v. Establish the most appropriate fairness metrics for the use case and document the rationale for selecting the particular fairness metrics.[15] Make sure to establish a cut-off or threshold for considering when to decommission the solution.

---

[14] The equity objectives may range from maintaining the current level of inequity to significantly reducing it.

[15] Choices about fairness metrics should be revisable based on the clinical evidence observed through real-time model use and an ongoing monitoring process (Mccradden M, Odusi O, Joshi S, et al. What's fair is… fair? Presenting JustEFAB, an ethical framework for operationalizing medical ethics and social justice in the integration of clinical machine learning. 2023 ACM Conf Fairness, Account, Transpar. Published online 2023:1505-1519. doi:10.1145/3593013.3594096).

### 4. Design AI solution workflow

[Assessment domain: Fairness, Fitness for purpose, Transparency]

Decision Point 4 describes how organizations design and test optimal workflows for clinician-facing AI products. It also explains how to adapt pre-existing operational structures, workflows, and technologies to enable successful integration of AI products. Decision Point 4 starts with understanding the current clinical workflow and ends with adapting the workflow to enable the implementation of the AI solution.

a. Ensure that the solution design is informed by meaningful and pragmatic recommendations from members of disadvantaged patient subgroups.
   [Active stakeholders: P] [Data source: Qualitative data]
   i. Recruit patients and patient advocates with lived experience as members of disadvantaged patient subgroups identified in 2(d)(iii).
   ii. Conduct interviews or focus groups with patients and patient advocates to surface problems and concerns they have for the solution design. Ensure to seek input from them about the various forms of support disadvantaged patient subgroups may need to benefit the most from using the solution.

b. Ensure that the solution design promotes inclusivity of clinical end-users and usability of the solution.
   [Active stakeholders: C] [Data source: Qualitative data]
   i. Recruit a representative sample of clinical end-users to test accessibility, inclusivity, and usability of the solution.
   ii. Conduct interviews, focus groups, or surveys with clinical end-users to surface user needs and concerns they have for the solution design. Ensure to seek input from them about the various forms of support they may need to use the solution efficiently.
   iii. Ensure to design the solution in a way that minimizes barriers to effective use for diverse clinical end-users.[16]

c. Design complementary non-technical solution components required to achieve equity objectives for implementation of the solution.
   [Active stakeholders: C, O, S]

[16] For example, issues like color blindness, sensitivity to light levels, and other ergonomic factors that may prevent end-users with disabilities from using the solution should be addressed (Mccradden M, Odusi O, Joshi S, et al. What's fair is… fair? Presenting JustEFAB, an ethical framework for operationalizing medical ethics and social justice in the integration of clinical machine learning. 2023 ACM Conf Fairness, Account, Transpar. Published online 2023:1505-1519. doi:10.1145/3593013.3594096).

i. Identify complementary non-technical solution components and resources that will be required to achieve each equity objective defined in 3(a)(ii).[17] Ensure to incorporate recommendations received in 4(a) and 4(b) into designing non-technical interventions.
ii. Evaluate internal resources and allocate necessary resources for implementing non-technical solution components.
iii. Develop training material and communication plan to equip frontline clinicians to achieve equity objectives defined in 3(a)(ii).

d. Align clinical end-users and organizational leaders to achieve equity objectives. [Active stakeholders: C, O, R, S]
   i. Communicate the equity objectives defined in 3(a)(ii) and their importance to organizational leaders. Ensure that the equity objectives are not presented as secondary to clinical or operational objectives.
   ii. Seek support from organizational leaders to implement necessary non-technical solution components defined in 4(c)(i).
   iii. Work with management and legal to establish incentives for frontline clinicians involved in the AI solution implementation.[18]

### 5. Generate evidence of safety, efficacy, and equity

[Assessment domain: Accountability, Fairness, Reliability and validity]

Decision Point 5 focuses on local validation of AI products before clinical use and identifying foreseeable and unresolved risks resulting from AI use in clinical care. It also describes how to determine if AI products should be clinically integrated or abandoned. Decision Point 5 starts with local validation of the AI solution and ends with a decision to integrate (or abandon) it in the clinical setting.

---

[17] Anticipate that AI solutions need to be complemented with other resources, workflows, and capabilities to effectively address inequities. Examples of non-technical solution components for the various categories of inequity may include:

- Ensuring sufficient capabilities, such as personnel and equipment, to perform diagnostic interventions or procedures for affected patient subgroups (workup inequity)
- Improving access to diagnostic testing and evaluation for affected patient subgroups (diagnosis inequity)
- Ensuring sufficient capabilities and access to treatments and interventions for affected patient subgroups (treatment inequity)
- Enhancing monitoring and sufficient capabilities to effectively manage affected patient subgroups (outcome inequity)

[18] Incentives may include public recognition for meeting performance targets, inclusion in promotion criteria, and possibly monetary awards under certain circumstances. Make sure that monetary awards related to effective use of AI do not induce or generate additional health service business, which may violate anti-kickback statutes.

a. Assess completeness and quality of local data required to construct model features across disadvantaged patient subgroups.
   [Active stakeholders: C, P, T] [Data source: Local healthcare retrospective data]
   i. Examine local healthcare retrospective data and assess missingness of all model features across disadvantaged and advantaged patient subgroups identified in 2(d)(iii).
   ii. Identify the model features that are missing at different rates for disadvantaged patient subgroups identified in 2(d)(iii).
   iii. For each model feature identified in 5(a)(ii), surface potential causes of differential missingness by consulting with a cross-functional team involved in data capture.
   iv. If the model features need to be sourced differently for disadvantaged patient subgroups, make necessary changes.
   v. Impute data in a fashion that minimizes inequities. If existing values within a disadvantaged patient subgroup identified in 2(d)(iii) differ substantially from values within an advantaged patient subgroup, consider imputation using subgroup statistics rather than full population statistics.
   vi. If missingness of a model feature continues to differ substantially for disadvantaged patient subgroups identified in 2(d)(iii), consider including a missing indicator as a model feature.

b. Seek an approval from an institutional review board, ethical review board, or research ethics board to access and use local healthcare prospective data.
   [Active stakeholders: R]

**For an AI solution that is being newly developed, proceed to the next set of procedures written in blue. For an AI solution that already exists, skip to 5(i).**

c. Analyze label choice bias across disadvantaged and advantaged patient subgroups.
   [Active stakeholders: C, O, P, T] [Data source: Training data]
   i. Examine a list of the actual labels used for model training.
   ii. From the list, select a closer-to-ideal label, even if it is available only for a small subset of patients. Use this label for analysis in the next procedures.
   iii. Consult with a cross-functional team and assess how well the actual label aligns with the ideal label identified in 2(d)(ii) across disadvantaged and advantaged patient subgroups identified in 2(d)(iii).
   iv. Ensure that the AI solution uses actual labels that accurately measure the ideal label. If the actual label does not accurately capture the ideal label, the solution has a risk of *label choice bias*.

d. Ensure that the model features are relevant to its actual label and capture the same meanings across disadvantaged and advantaged patient subgroups.
   [Active stakeholders: C, T] [Data source: Training data]

i. Examine the actual labels and a comprehensive list of model features used for model training.
ii. Ensure that all model features accurately represent concepts relevant to the actual label.
iii. Consult with clinicians and understand if there are known differences in how model features are gathered and represented across disadvantaged patient subgroups identified in 2(d)(iii).[19]
iv. For each feature, examine how it is correlated with actual labels across all patient subgroups and identify whether this association manifests inconsistently in disadvantaged patient subgroups identified in 2(d)(iii).
v. Ensure that the AI solution uses features that have consistent meanings and are captured robustly across all patient subgroups. If the association between model features and the actual label is inconsistent across different patient subgroups, the solution has a risk of *measurement bias*.

e. Identify potential hidden stratification that masks unequal model performance between disadvantaged and advantaged patient subgroups.
[Active stakeholders: T] [Data source: Training data]
i. Identify diagnostic and treatment subgroups within the actual label.[20]
ii. Compare model performance between the identified subgroups.
iii. If the model performance of a subgroup with diagnosis and treatment is better than the model performance of a subgroup without diagnosis and treatment, flag for *hidden stratification*.[21] Consider alternative solutions (return to 2(a)) or retrain the model after excluding a cohort of patients from the subgroup with diagnosis and treatment.
iv. If the model performance of a subgroup without diagnosis and treatment is better than the model performance of a subgroup with diagnosis and treatment, or if model performance between the subgroups is similar, then compare model performance between disadvantaged and advantaged patient subgroups defined in 2(d)(iii) within each diagnostic and treatment subgroup.
v. Ensure that the AI solution performs equally well on disadvantaged and advantaged patient subgroups within each diagnostic and treatment subgroup.

[19] For example, examine whether there are any known symptoms that manifest differently for different patient demographics.
[20] For example, diagnostic and treatment subgroups may include (1) a subgroup that has completed all relevant prior workup versus a subgroup who has not (workup inequity), (2) a subgroup that is diagnosed at an earlier disease state versus a subgroup that is diagnosed at a later disease state (diagnosis inequity), (3) a subgroup that receives important interventions prior to or immediately after diagnosis versus a subgroup that does not receive important interventions (treatment inequity), and (4) a subgroup with poor outcomes versus a subgroup with good outcomes (outcome inequity).
[21] Oakden-Rayner L, Dunnmon J, Carneiro G, Ré C. Hidden stratification causes clinically meaningful failures in machine learning for medical imaging. In: Vol 52. ACM CHIL '20: ACM Conference on Health, Inference, and Learning. ; 2020:151-159. doi:10.1145/3368555.3384468

f. Assess model performance and compare it between disadvantaged and advantaged patient subgroups.
[Active stakeholders: T] [Data source: Training data]
   i. Assess model performance for disadvantaged patient subgroups and advantaged patient subgroups identified in 2(d)(iii).
   ii. Compare the model performance of disadvantaged patient subgroups to the model performance of advantaged patient subgroups.
   iii. If model performance is worse for disadvantaged patient subgroups than model performance for advantaged patient subgroups, the implication depends on whether health inequities were identified in 2(d)(ii):
      ▪ If health inequities were confirmed, the difference in model performance could be due to existing health inequities, not due to modeling, and thus, conveys minimal concern with the model.
      ▪ If health inequities were not confirmed, the difference in model performance could be due to modeling and create future health inequities and thus, conveys potential concern with the model.
   iv. If model performance for disadvantaged patient subgroups is similar to model performance for advantaged subgroups, the implication depends on whether health inequities were identified in 2(d)(ii):
      ▪ If health inequities were confirmed, the similarity in model performance could be due to modeling and further reinforce existing health inequities and thus, conveys potential concern with the model.
      ▪ If health inequities were not confirmed, the similarity in model performance could be due to the absence of health inequities and thus, conveys minimal concern with the model.
   v. If the model has potential concern, retrain the model to ensure that the model has minimal concern.[22]

g. Determine which SDOH and demographic data are appropriate to be included in the model to minimize potential risk of worsening health inequities.
[Active stakeholders: T] [Data source: Training data]
   i. Review the comprehensive list of model features gathered in 5(d)(i).
   ii. If the solution uses patient SDOH and demographic data as model features, gather a rationale for inclusion of each SDOH and demographic feature.
   iii. Assess model performance without using SDOH and demographic data as model features.
   iv. Compare model performance data measured in the previous procedure 5(g)(iii) with model performance data measured in 5(f)(i) across disadvantaged and advantaged patient subgroups identified in 2(d)(iii).[23]

---

[22] For patient subgroups identified in section 2(d)(iii) who are poorly represented in the local context (e.g., Native American women in Boston), AI solution performance may have wide confidence bounds. Ensure that performance measures account for uncertainty.
[23] Sometimes, removing SDOH and demographic data as model features may decrease model performance. For example, removing race worsened model performance (Khor S, Haupt EC, Hahn EE,

v. Use SDOH and demographic data to improve performance for disadvantaged patient subgroups identified in 2(d)(iii). However, if model performance remains robust across disadvantaged patient subgroups without using SDOH and demographic data, minimize use of this data.

h. Assess prospective model performance and compare it to retrospective model performance across disadvantaged and advantaged patient subgroups.
[Active stakeholders: T] [Data source: Training data, Local healthcare prospective data]
    i. Conduct retrospective analysis of model performance on local healthcare retrospective data across disadvantaged and advantaged patient subgroups identified in 2(d)(iii) against fairness metrics identified in 3(b)(v).
    ii. Conduct prospective analysis of model performance on local healthcare prospective data across disadvantaged and advantaged patient subgroups identified in 2(d)(iii) against fairness metrics identified in 3(b)(v).
    iii. Ensure that model performance across disadvantaged and advantaged patient subgroups identified in 2(d)(iii) is consistent across local retrospective and local prospective settings against fairness metrics identified in 3(b)(v).

**For an AI solution that is being newly developed, skip to 5(j).**

i. Perform comprehensive assessment of model performance across disadvantaged and advantaged patient subgroups.
[Active stakeholders: T] [Data source: Local healthcare retrospective data, Local healthcare prospective data, Training data]
    i. Conduct retrospective analysis of model performance on local healthcare retrospective data across disadvantaged and advantaged patient subgroups identified in 2(d)(iii) against fairness metrics identified in 3(b)(v).
    ii. Conduct prospective analysis of model performance on local healthcare prospective data across disadvantaged and advantaged patient subgroups identified in 2(d)(iii) against fairness metrics identified in 3(b)(v).
    iii. Compare model performance on retrospective data derived in 5(i)(i) and model performance on prospective data derived in 5(i)(ii) to model performance data gathered from external settings in 2(f)(i).
    iv. Ensure that model performance across disadvantaged and advantaged patient subgroups identified in 2(d)(iii) is consistent across local retrospective, local prospective, and external settings against fairness metrics identified in 3(b)(v).

j. If the model performs worse for a certain patient subgroup on local prospective healthcare data, consider adapting the model or its use to help minimize negative impacts on inequities.
[Active stakeholders: T] [Data source: Local healthcare retrospective data]

---

Lyons LJL, Shankaran V, Bansal A. Racial and Ethnic Bias in Risk Prediction Models for Colorectal Cancer Recurrence When Race and Ethnicity Are Omitted as Predictors. JAMA Netw Open. 2023;6(6):e2318495. doi:10.1001/jamanetworkopen.2023.18495).

i. Consider the following to improve model performance for the patient subgroups negatively affected by poor model performance:
   - Oversample patients from the affected subgroup to build the model.
   - Source additional data and features that are specifically important for the affected patient subgroup.
   - Process data differently, including different approaches to imputing data, that are specific to the affected patient subgroup.
   - Build a separate model specifically targeted for the affected patient subgroup.
   - Change the model threshold for the affected patient subgroup. Assess whether a different threshold could achieve similar levels of sensitivity, even if positive predictive value or precision suffers.[24]

ii. Consider the following if model performance for a certain patient subgroup cannot be improved:
   - Narrow the scope of use of the model to only be used on patient subgroups for whom equity objectives defined in 3(a)(ii) are attainable. Note that it is possible that a model that performs poorly on disadvantaged patient subgroups can still achieve equity objectives.[25]
   - Consider alternative, non-AI solutions (return to 2(a)).

k. Ensure that model performance aligns with the equity objectives.
[Active stakeholders: T] [Data source: Local healthcare retrospective data, Local healthcare prospective data]

i. Ensure that model performance measures derived in 5(h)(iii) or 5(i)(iv) align with the requirements for achieving the equity objectives defined in 3(a)(ii).[26]

ii. If model performance does not align with equity objectives defined in 3(a)(ii), consider either selecting an alternate solution (return to 2(a)) or adapting the model (return to 3(a)).

l. Conduct a prospective pilot study to validate whether the AI solution achieves equity objectives.
[Active stakeholders: C, P, T] [Data source: Local healthcare prospective data, Qualitative data]

---

[24] In some cases (e.g., targeted vaccination campaigns and equity efforts in doulas), it may be appropriate to provide unequal resourcing and intervention to different groups of patients to close the potential inequities.

[25] For example, imagine that there is a model built with an intention to de-escalate an intervention that currently harms Black patients at a higher rate than White patients. The model is found to be less accurate on Black patients than White patients. However, as long as the model identifies some Black patients, differences in model performance should not be too concerning because it is still better than the status quo.

[26] For example, a model exhibiting minimal lead time (i.e., model identifies the outcome of interest shortly before the event occurs) would be ineffective at addressing a diagnostic inequity (i.e., more severe disease progression at the time of diagnosis).

i. Recruit a diverse sample of clinical end-users and patients. Ensure that disadvantaged patient subgroups identified in 2(d)(iii) are adequately represented in the pilot study context.
ii. Randomly assign units, frontline clinicians, or individual patients to control (baseline approach without the new AI solution) and intervention (with the new AI solution) groups. If unable to conduct a randomized study, conduct an observational study that controls for confounders, such as interrupted time series or differences-in-differences study designs.
iii. Couple quantitative assessment of patient outcomes with qualitative analyses of frontline clinicians and patients. Ensure that patient representatives from disadvantaged patient subgroups identified in 2(d)(iii) are included in qualitative research efforts.
iv. Assess performance of the model according to fairness metrics identified in 3(b)(iv) during the prospective pilot study.
v. Assess the degree of adoption and effectiveness of non-technical solution components defined in 4(c)(i).
vi. Assess measurably progress towards achieving equity objectives specified in 3(a)(ii).
vii. Report results of 5(l)(iv), 5(l)(v), and 5(l)(vi) to frontline clinicians and local patient community members.

### 6. Execute AI solution rollout

[Assessment domain: Accountability, Fairness, Transparency]

Decision Point 6 describes how to disseminate information about AI solutions to affected clinicians, including end-users, and manage changes in the workflow caused by clinical integration of AI solutions. It also explains how to prevent misuse of AI solutions beyond their intended scope. Decision Point 6 starts with a clinical integration of the AI solution and ends with a decision to continue using the solution in the clinical setting.

a. Document information about the model development and implementation and share it with clinical end-users, members of disadvantaged patient subgroups, and others who may be affected by use of the model.
[Active stakeholders: C, O, P, T]
    i. Create communication material with the following information:
        - Equity objectives defined in 3(a)(ii)
        - Fairness metrics identified in 3(b)(v)
        - Information about non-technical solution components defined in 4(c)(i) meant to support achieving the equity objectives
        - Model performance across disadvantaged patient subgroups identified in 5(h)(iii) or 5(i)(iv)
        - Implementation domain, directions, workflow, and warnings

ii. Tailor the communication materials to be understandable and relevant to clinical end-users, members of disadvantaged patient subgroups identified in 2(d)(iii), and others who may be affected by use of the model.
iii. Secure approval of communication materials for sharing.
iv. Develop a communication plan to share the materials with the personnel identified in 6(a)(ii).
v. Operationalize the communication plan and ensure the materials reach the personnel identified in 6(a)(ii).

b. Educate clinical end-users about potential bias in using the solution. [Active stakeholders: C]
   i. Make clinical end-users aware of baseline inequities surfaced in 2(d)(ii) and the equity objectives set in 3(a)(ii).
   ii. Educate clinical end-users about confirmation bias and its harmful consequences on health inequity.
   iii. Provide clinical end-users direct and explicit instructions on what they are supposed to do with the model output and any specific actions they need to take to achieve equity objectives set in 3(a)(ii).[27]

c. Where applicable, seek an approval from an institutional review board, ethical review board, or research ethics board to implement and use the AI solution in clinical practice. [Active stakeholders: R]

d. After rollout, continue to seek feedback from clinical end-users and members of disadvantaged and advantaged patient subgroups to achieve equity objectives. [Active stakeholders: C, P, T] [Data source: Qualitative data]
   i. Review recommendations surfaced in 4(a) and the non-technical solution components defined in 4(c)(i).
   ii. Create a communication plan to seek feedback on the solution and surface unanticipated challenges associated with the solution from clinical end-users and members of disadvantaged and advantaged patient subgroups identified in 2(d)(iii).
   iii. Seek feedback from clinical end-users and members of disadvantaged and advantaged patient subgroups identified in 2(d)(iii) on how well the solution progresses toward achieving equity objectives defined in 3(a)(ii).
   iv. If challenges persist and limit progress towards achieving equity objectives, consider updating the AI solution or workflow (see Decision Point 8).

[27] Meanwhile, it is important to respect clinician autonomy.

#### 7. Monitor the AI solution

[Assessment domain: Fairness, Reliability and validity, Transparency]

Decision Point 7 focuses on monitoring AI solutions and affected workflow over time and sustaining improved outcomes achieved through AI use. It also highlights the importance of conducting audits of AI products that complement ongoing monitoring and proactively identifying risks not envisioned at the time of clinical integration.

a. Regularly monitor the model performance across disadvantaged and advantaged patient subgroups.
   [Active stakeholders: P, T] [Data source: Local healthcare prospective data]
   i. Monitor model performance across disadvantaged and advantaged patient subgroups identified in 2(d)(iii).
   ii. Monitor fairness metrics identified in 3(b)(iv).
   iii. Share monitoring outcomes with clinical end-users, members of disadvantaged and advantaged patient subgroups identified in 2(d)(iii), and others who may be affected by use of the model via pre-established communication plan in 6(d)(ii).

b. Regularly monitor the work environment of the solution across disadvantaged and advantaged patient subgroups.
   [Active stakeholders: O, P, T] [Data source: Qualitative data]
   i. Monitor progress towards achieving equity objectives specified in 3(a)(ii).
   ii. Monitor the degree of adoption of non-technical solution components defined in 4(c)(i).
   iii. Share monitoring outcomes with clinical end-users, members of disadvantaged and advantaged patient subgroups identified in 2(d)(iii), and others who may be affected by use of the model through the pre-established communication plan.
   iv. Seek feedback from clinical end-users, members of disadvantaged and advantaged patient subgroups identified in 2(d)(iii), and others who may be affected by use of the model to understand their experience with the AI solution and progress towards achieving equity objectives.

c. Regularly monitor health inequities across disadvantaged and advantaged patient subgroups.
   [Active stakeholders: C, O, P, T] [Data source: Local healthcare prospective data]
   i. Calculate health inequities identified in 2(d)(ii) for disadvantaged and advantaged patient subgroups identified in 2(d)(iii).
   ii. Compare the level of inequities calculated in 7(c)(i) with the baseline level calculated in 2(d)(ii).
   iii. Ensure that any changes in the level of inequities align with equity objectives defined in 3(a)(ii).

iv. Consult with frontline clinicians, management, and members of disadvantaged patient subgroups identified in 2(d)(iii) on an ongoing basis and validate whether the quantitative measures reflect reality on the ground.
v. If inequities worsen in ways that were not anticipated and undermine equity objectives, proceed to 8(a) and 8(b).[28]

### 8. Update or decommission the AI solution

[Assessment domains: Accountability, Fairness, Reliability and validity, Transparency]

Decision Point 8 focuses on updating or decommissioning AI products, adapting work environments affected by AI use, and expanding the use of AI products to new settings. It also describes how to minimize disruptions and eliminate harms that result from decommissioning AI products and disseminating information about AI product updates to affected clinicians.

a. Determine updates to the model or its work environment.
[Active stakeholders: C, O, P, T] [Data source: Local healthcare prospective data]
    i. If metrics monitored in 7(a)(i) and 7(a)(ii) reveal changes, engage a cross-functional team to determine the need for updates.
    ii. When changes in model performance across disadvantaged patient subgroups identified in 2(d)(iii) or changes in fairness metrics identified in 3(b)(iv) are observed, consider executing activities described in 5(j)(i) to improve model performance for the disadvantaged patient subgroups.
    iii. When changes in the effectiveness of non-technical solution components defined in 4(c)(i) or changes in progress toward achieving equity objectives defined in 3(a)(ii) are observed, reassess model performance as well as non-technical solution components defined in 4(c)(i).[29] Consider revisiting 4(a) and 4(c) to update non-technical interventions that can better achieve equity objectives.
    iv. When changes in the model or workflow are made, ensure to communicate information about the changes to all who may be affected by the changes through the pre-established communication plan.

b. If updating the model or its work environment does not improve model performance or fails to improve progress towards equity objectives, consider decommissioning the model.
[Active stakeholders: C, O, P]
    i. Revisit potential solutions identified in 2(a) and consider alternative solutions that are well equipped to improve health equity for disadvantaged patient subgroups identified in 2(d)(iii).

[28] Note that failing to meet equity objectives may be due to ineffective implementation or adoption of non-technical solution components defined in 4(c)(i) rather than inequities in AI model performance.
[29] For example, there may be insufficient staffing or capacity available to respond to AI model outputs to address inequities.

ii. Conduct a final assessment of the model and report outcomes to both frontline clinicians and members of disadvantaged patient subgroups identified in 2(d)(iii).
iii. Consider decommissioning the model, if an alternative solution is better equipped to improve health equity for disadvantaged patient subgroups or the results of the final assessment are poor based on decommissioning cut-offs established in 3(b)(iv) and 3(a)(ii).
iv. When a decision to decommission the model is made, ensure to communicate information about the decision to all who may be affected by the decision through the pre-established communication plan.

c. If the model successfully achieved equity objectives and there is interest to expand model use, evaluate appropriate technical and non-technical resources to expand the AI solution to new settings.
[Active stakeholders: C, O, P] [Data source: Local healthcare retrospective data, Local healthcare prospective data]]
    i. Assess baseline health inequities in the new implementation context, following a procedure described in 2(d)(ii).
    ii. Assess model performance across disadvantaged patient subgroups identified in 2(d)(iii) in the new implementation context.
    iii. Define equity objectives for the new implementation context, following a procedure described in 3(a)(ii).
    iv. Design non-technical solution components to help achieve equity objectives following procedures described in 4(c) in the new implementation context.
    v. Assess model performance following procedures described in 5(f) in the new implementation context.
    vi. Conduct a prospective pilot study following procedures described in 5(l) in the new implementation context.

#### Glossary

**Actual label.** A.k.a. *actual target*. Downstream outcome used to train or evaluate a model.

**Advantaged patient group.** A group of patients who are least likely to be negatively impacted by health inequities.

**Baseline population.** Population in a geographic setting where a healthcare delivery organization is based.

**Computable ideal label.** A computable proxy for an ideal label that can be derived from structured or unstructured data elements available within local data.

**Confirmation bias.** The tendency to gather evidence that confirms preexisting expectations, typically by emphasizing or pursuing supporting evidence, while dismissing or failing to seek contradictory evidence.

**Cross-functional team.** A team composed of personnel with a variety of expertise who come together to achieve a common goal. In this framework, a cross-functional team is composed of all active stakeholders listed in each procedure.

**Disadvantaged patient group.** A group of patients who are most likely to be negatively impacted by health inequities.

**End-user.** A person who uses and interacts with the product. Typically, end-users are frontline clinicians.

**Feature.** Data in its final form post cleaning and QA that is being fed directly into model training and evaluation or for model inference.

**Hidden stratification.** The data contains unrecognized subsets of cases which may affect model training, measured model performance, and most importantly the clinical outcomes related to the use of a medical image analysis system.

**Ideal label.** A.k.a. *ideal target* or *ground truth outcome*. Clinical outcome used for model training that is validated by clinicians when pristine and complete information about patients is available for model development.

**Label choice bias.** A mismatch between the ideal label and the actual label that results in worse model performance for a disadvantaged patient group.

**Local healthcare retrospective data.** Historical healthcare data that is curated within the primary healthcare delivery organization seeking to adopt an AI product. The local data can be sourced from a variety of systems, including the EHR, radiology PACS system, medical claims, audit logs, electrocardiograms, and high-frequency vital sign monitors. When a model is internally developed, the local healthcare retrospective data set is used for training the model.

**Local healthcare prospective data.** Real-time healthcare data that is curated within the primary healthcare delivery organization seeking to adopt an AI product. The local data can be sourced from a variety of systems, including the EHR, radiology PACS system, medical claims, audit logs, electrocardiograms, and high-frequency vital sign monitors. The local healthcare prospective data set is used for validating a model during a ‘silent trial’ and for using the model in clinical care.

**Local non-healthcare data.** Non-healthcare data that is curated within a geographic setting where a healthcare delivery organization is based. The local non-healthcare data can be derived from a variety of external sources, including US Census.

**Missing indicator.** A binary variable that indicates whether a model feature is missing or not.

**Patient group.** A collection of patients with similar experiences, cultural norms, practices, or way of life.

**Representation bias.** A mismatch between the demographic makeup of the local population, especially regarding disadvantaged patient groups, and the demographic makeup of the training data.

**Training data.** Data used for training a model. When the model is externally developed, the training data set contains data from an external source. When the model is internally developed, the training data set is sourced from local healthcare retrospective data.

**Work environment.** A sociotechnical environment where an AI solution is being used and interacted by its end-users and others who are affected by its use.
